# The Mortality Burden from COVID in Low-Income Settings: Evidence from Verbal Autopsies in India

**DOI:** 10.1101/2024.01.02.24300728

**Authors:** Anup Malani, Wanran Zhao

## Abstract

Measuring the mortality burden of SARS-CoV-2 in lower-income countries is difficult because death registries are incomplete and lack cause of death. We address this problem in India, which had the second-highest number of officially reported cases. We completed WHO-compliant verbal autopsy (VA) surveys on roughly 20,000 deaths drawn from a population-representative sample. SARS-CoV-2 deaths spike in June 2020, just after India’s lockdown, and in May 2021, after its second wave. During those spikes the virus is responsible for 23.3% and 35.8% of all deaths, respectively. Cardiovascular deaths also spike during the start of the pandemic. We find that the death rate rises by 81% during the pandemic, SARS-CoV-2 is responsible for 33% of these excess deaths, and cardiovascular disease for 23% of these deaths.

## I. Introduction

Even as the COVID pandemic has eased, it is important to evaluate its impact. Measuring its health impact more precisely is critical for determining both the effectiveness and cost-effectiveness of policy responses, including non-pharmaceutical interventions such as lockdowns and school closures and medical interventions such as vaccines. Estimating the mortality burden of SARS-CoV-2, however, is no easy task. In higher-income countries, where there are well-developed reporting systems, there remain debates about causation ^1,2^. In lower-income countries, where such systems are underdeveloped or missing, the challenges are greater still ^3^. India is a case in point.

India is among the countries that were hit the hardest by the COVID pandemic. Through 2021, it reported the second highest-number of confirmed SARS-CoV-2 cases (∼44 million) and third-highest number of SARS-CoV-2 deaths (∼530,000) of all countries ^4^. Even these high numbers might underestimate the virus’s impact on the country. Estimates of excess deaths–the increase in all-cause deaths from 2020 to mid-2021 relative to baseline–range from 4 to 6 million through mid 2021 ^5–9^.

These excess mortality estimates are contested, however, because India has poor measures of deaths specifically attributable to SARS-CoV-2. Estimates of excess deaths are typically based on India’s state Civil Registration System (CRS). While those collect data on deaths, they do not attribute a cause to deaths ^10^. (Non-official surveys that report deaths have similar problems^5,6^. Therefore, they may over-estimate deaths attributable directly to SARS-CoV-2. During the pandemic, India began to report the number of decedents who tested positive for SARS-CoV-2 or were presumed to be SARS-CoV-2 positive. These are the basis of official COVID death reports. However, these reports may underestimate SARS-CoV-2 deaths because a small percentage of decedents were tested or diagnosed for SARS-CoV-2 ^11^. India’s Sample Registration System (SRS), which samples a random 1% of the population, conducts verbal autopsies to determine cause of death ^10^. However, the SRS for 2020-2021 has not yet been completed.

To determine how much of excess mortality is attributable to SARS-CoV-2 as opposed to the pandemic overall, we obtain a representative sample of decedents in India and ascertain cause of deaths using WHO-compliant verbal autopsy surveys administered to next-of-kin. The representative sample was obtained from the Consumer Pyramids Household Survey and contained all deaths reported in that survey between 2018-2021. The verbal autopsies revealed cause-of-death encoded in 3-digit ICD10 codes. This exercise is intended not merely to obtain better estimates of SARS-CoV-2 attributable deaths in India, but to explore the value of ex post verbal autopsies as a method of measuring the burden of the pandemic in lower-income countries.

## II. Methods

### Funding and Approvals

The verbal autopsy survey on which this study is based was commissioned by Emergent Ventures, and conducted by the Centre for Monitoring the Indian Economy (CMIE), a commercial enterprise. Emergent Ventures requires that the data from the survey, without person identifiers, to be released to the public. CMIE also conducts a household demographic and economic survey, the Consumer Pyramids Household Survey (CPHS). CPHS was the sampling frame for our verbal autopsy survey. The latter also includes identifiers that allow users to merge it with data from CPHS, which can be purchased for a subscription fee by any member of the public.

The authors of this paper served as unpaid advisors to CMIE for designing the sample frame and vetting the verbal autopsy survey, but did not have access to any personally identifiable information. They received the verbal autopsy data for free, before it is released to the public. The Becker-Friedman Institute paid for a University of Chicago subscription to the de-identified CPHS data.

The University of Chicago IRB determined this study (Case No. IRB22-0119) was not human subjects research as no identifiable information is shared and the data are or will be publicly accessible. Prabhat Jha and Leslie Newcombe, who managed the physician panel that mapped map e-VA results to ICD10 codes at the Centre for Global Health Research, have umbrella approval to provide broad support for the creation of mortality datasets via verbal autopsies from the IRB of Unity Health hospital system (Case no. REB #15-231).

### Sample and data

As a first step, we create two samples: a representative sample of the Indian population alive at some point from 2018-2021 and a representative sample of Indians who died between 2018-2021. The first sample serves as a denominator and the second sample as a numerator in the estimation of mortality rates. The latter also serves as a sample frame for verbal autopsies.

Both living and deceased samples are drawn, in part, from a panel survey called the Consumer Pyramids Households Survey (CPHS), produced by the Centre for Monitoring Indian Economy (CMIE), Mumbai, India. This is a panel data set on 236,908 unique households representative of urban and rural areas of all states except those in the northeast of India (Vyas 2020). The CPHS attempts to survey each household every 4 months (a round), with a representative quarter of sample households surveyed each month. The survey spans the period 2014-2022. While the CPHS is primarily used to measure the income and consumption of household members, it also updates household rosters in each round, allowing one to track those who are living and deceased. Supplement Section I provides more details on the CPHS survey.

Our representative sample of living Indians is the set of all living persons surveyed by CPHS between 2018-2021, a period that covers 2 years before the pandemic and 2 years during it.

Ideally, our representative sample of deceased persons would include all persons who would otherwise be in the living sample during the same subset of years (2018-2021), but who died in that period. However, our actual representative sample of deceased persons is a subset of this ideal representative sample. It is a subset because the CPHS data have three problems. <u>First</u>, these data do not report the cause of death. <u>Second</u>, they are over-inclusive: some deaths reported in 2018-2021 actually occurred before 2018. The CPHS surveys households every 4 months and only asks if a household member died, not when they died. Therefore, a household surveyed in January - April 2018 might report a death that occurred in September - December 2017. <u>Third</u>, they are under-inclusive: some deaths that occurred in 2018-2021 are not reported in CPHS until after 2021. For example, if a household is surveyed by CPHS in September 2021 but experienced a death in October - December 2021, that death would not be reported to CPHS until January 2022 and therefore omitted from our sample. The latter two problems are compounded if a household skips a CPHS round.

To address the cause-of-death and over-inclusive deaths problems, CMIE conducted WHO-sanctioned verbal autopsy (VA) surveys on all deaths reported on CPHS rosters from 2018-2021. These VA surveys asked the next-of-kin in each deceased CPHS sample member’s household the date of and circumstances of the decedent’s passing. Cause-of-death was determined by a panel of doctors based on answers to a range of questions concerning the circumstances of the decedent’s death. We address the over-inclusive problem by dropping all deaths that occurred before 2018.

The under-inclusive problems are addressed with a statistical “under-inclusivity adjustment” explained below, in the section on “Weights for deceased persons.”

### Verbal autopsy survey and cause of death

CMIE conducted in-person visits to each household that reported a death in the CPHS survey during 2018-2021. At each household, relatives of the deceased person were asked to complete an electronic verbal autopsy (e-VA) survey. The e-VA employed the 2016 WHO Verbal Autopsy Standard tool, version 1.5.1.,^12^ for neonates, children, adults, and stillbirths. The main change is that a few questions are added to identify symptoms associated with SARS-CoV-2. The e-VA software was run on laptops. Data were validated as they were entered; random re-sampling was employed for quality control. Details on the instrument are provided in a prior publication.^13^

Each death was assigned an International Classification of Diseases (tenth edition, ICD10) code that we treat as the cause of death by the Centre for Global Health Research, under the oversight of Prabhat Jha and Leslie Newcombe. The WHO added 3 codes to the ICD-10 classification to capture SARS-Cov-2 in 2021: U07.1, U07.2 and U08 (https://icd.who.int/browse10/2019/en<u>#/</u>). This assignment was done by two physicians on the basis of their review of the e-VA answers for the decedent. The two physicians we selected from a panel of physicians specially trained on death certification and ICD coding of verbal autopsies. Following the same procedure as the Indian Million Death Survey,^14,15^ these physicians coded deaths independently and anonymously, and a third physician adjudicated any differences between the coding of the first two physicians, whose identities were masked.

We aggregate causes of deaths (aside from SARS-CoV-2) using a standard death classification from the Indian National Burden Estimates^14^ and aligned with the WHO Global Health Estimates.^16^ We classify deaths in our sample as caused by SARS-CoV-2 in 2 steps. First, if the 2-digit ICD10 code assigned to a decedent from the VA survey is U07 or U08, we call it a “potential SARS-CoV-2” death. This assignment is over-inclusive because U08 includes non-SARS-CoV-2 deaths (e.g., vaping-related disorders). Moreover, there may be errors in the verbal autopsies coding of cause of death, causing false positives (non-SARS-CoV-2 deaths coded as SARS-CoV-2) and false negatives (SARS-CoV-2 deaths reported as non-SARS-CoV-2). Second, to address either cause of over-inclusion, we calculate “SARS-CoV-2-attributable” excess deaths by subtracting the average annual rates of potential SARS-CoV-2 deaths in 2018-2019 from the average annual rates of those deaths in 2020-2021.

### Outcomes

Our primary outcomes include the number of deaths and death rate attributable to all causes and to SARS-CoV-2, the top 5 other cause of death, and to other causes, both in our completed VA survey sample and in the population. Our primary outcomes also include the excess number of deaths attributable to all causes and to these same specific causes during the pandemic in the population. Our secondary outcomes are the these the primary outcomes, but for specific subpopulations: 5 different age groups, males and females, rural and urban residents, and households in each of 4 income quartiles.

### Dates and date of death

We count time in months. We define the pandemic to be the period from January 2020 to December 2021. India’s national lockdown, which ran from March 24 to May 31, 2020, is coded as occurring during April and May 2020. Graphs mark September 2020 and April 2021 as the peak of India’s first and second COVID waves based on peaks in officially reported infections ^4^.

The date of death is obtained in two steps. The CPHS provides the date that a household was surveyed and *reported* a member (the decedent) as having died since the last CPHS survey. This CPHS report date is later than the date of death because CPHS surveys households every 4 months and asks *if* a household member died, not when they died. Therefore, our first step is that CMIE, while conducting the verbal autopsy or a follow-up phone survey, asked each decedent’s family the specific date of the decedent’s death. In 1,569 cases, the household incorrectly remembered the decedent’s date of death and gave a date *after* the CPHS report date, which is not possible. Our second step is that, in these 1,569 cases, we list the month of death as the month that is the midpoint between the (a) first month that the decedent’s household answered a CPHS survey and reported the decedent as having passed and (b) the last month that households answered a CPHS survey and did not report that the decedent had passed. We call this date the midpoint date of death.

In the Supplement, we report an alternate set of results that equates the date of death for each decedent with the midpoint date of death, ignoring the specific dates households gave during the VA survey.

### Statistical methods

The death rate from SARS-CoV-2 in a period is defined as the number of people who died from the disease (numerator) divided by the number of living people in that period plus the number of people who died in that period (denominator). The death rate may be unweighted, in which case it provides estimates about the sample. If it is weighted appropriately, it can provide estimates about the population. Using this framework, we provide information on the inputs into estimates of the death rate.

### Weights for living persons

To extrapolate from our representative sample to the whole population of India, we need sampling weights. For each living person in our sample, CPHS provides weights are a composite of inverse probability sampling weights and adjustments for non-response in specific survey rounds ^17^. Details on these weights for the living population are provided in the Supplement. These “CPHS weights” permit the estimation of summary statistics that are unbiased for the living Indian population at the national, rural, and urban levels for each month. We also sum these “CPHS weights”, which determine the number of people each living respondent represents, across all living CPHS respondents in each month to estimate the total living population in each month.

### Weights for deceased persons

There are three hurdles to the generation of weights for decedents in our sample. First, CPHS does not provide such weights for deceased sample members for the month in which they are reported to CPHS as having died. To address this problem, we impute “CPHS weights” for decedents in one of two ways. One way is to assign the decedent a weight equal to the average CPHS weight of living members in the decedent’s household in the month of death. If the decedent has no other household members surveyed that month, a second way is to assign them the average CPHS weight of other living respondents in CPHS that month. We use these imputed deceased weights to calculate population-level means for and totals of deceased persons.

The second hurdle is that, because households may be surveyed by the CPHS months after a member has died, our sample of deaths reported to the CPHS may miss deaths that occurred before 2022 but had not been reported to the CPHS by the end of 2021. We address this problem in two steps. Step one is to use the first 16 months of the VA survey sample to estimate the probability p(t) that a death in a given month is reported to CPHS within t months, where t ranges from 1 to 32. Step two is, for each death that is reported in t months before December 2022, we create a scaling-up factor, equal to 1/p(t). This scaling-up factor, which we call an “under-inclusivity adjustment”, can be interpreted as the number of actual CPHS sample deaths that each reported CPHS death represents. Additional details on this adjustment are provided in the Supplement.

The third hurdle is that not all CPHS sample households with a reported decedent respond to our VA survey. Moreover, those who do respond may not be representative of those who do respond to the VA. To address this non-random undercounting of the numerator due to non-response to the VA, we compute an adjustment in three steps. First, we estimate a prediction model for whether a household with a decedent reported to the CPHS responds to the VA survey. This prediction model includes household demographic and economic covariates, as well as strata fixed effects and the CPHS weights. Second, we create a propensity-score weight for each decedent whose household responded to the VA survey. This “VA non-response” weight is equal to the reciprocal of the predicted probability that the decedent’s household responds to the verbal autopsy. Additional details on this weight are provided in the Supplement.

Our composite weight for all decedents who respond to the VA survey is the product of the CPHS weight, the under-inclusivity adjustment, and the VA non-response weight.

### Count of living persons

The denominator includes a count of living persons in the CPHS sample. The <u>unweighted count</u> <u>of all living persons</u> in a month is the set of all living persons whose households responded to a CPHS survey in that month. The <u>response rate</u> for the CPHS survey in a month is the fraction of households that CPHS attempted to survey in a month that responded to that survey in that month. Our estimate of <u>the number of all living persons in the population</u> in a month is the sum of the weights of all persons who responded to the CPHS in that month. These weights sum up to the number of individuals the CPHS projects to be in the population in a given month.

### Count of deaths

The <u>unweighted count of all-cause or cause-specific VA deaths</u> in a month is estimated with the total number of decedents in the actual VA sample who died from the designated cause(s) and whose date of death is that month. We estimate the <u>all-cause or cause-specific number of</u> <u>deaths in the CPHS sample</u> by calculating the weighted sum of deaths reported in the VA sample from the designated cause(s) in that month, where the weight on each death is the product of the associated VA non-response weight and the under-inclusivity adjustment. We estimate <u>the number of all-cause or cause-specific deaths in the population</u> with a similar weighted sum, except that each weight is the product of the VA non-response weight, the under-inclusivity adjustment, and the imputed CPHS weight for a decedent.

### Count of excess deaths

The number of <u>all-cause or cause-specific excess deaths in the CPHS sample</u> in a month during the pandemic is the all-cause or cause-specific number of deaths in the CPHS sample in that month minus the average number of such deaths each month during 2018-2019.

### Death rate

The <u>unadjusted all-cause death rate in the CPHS sample</u> in a period is estimated with the fraction of the CPHS sample that dies during that period. If the period is, for example, a month, the numerator is the number of deaths in the VA sample in a month and the denominator is the number of CPHS living respondents in a month. Note that CPHS sample members who die in that month are not included in the denominator, producing a slight overestimate of the all-cause death rate in the sample. We discuss this problem further when explaining our estimates of population-level death rates below.

The <u>VA non-response adjusted death rate</u> modifies the numerator to be the sum of non-response adjustments for each decedent in a month. The <u>under-inclusivity adjusted death rate</u> modifies the numerator further: it is the product of each decedent’s VA non-response weight and under-inclusivity adjustment, summed over all VA decedents in a month.

Our estimate of <u>the death rate in the population</u> modifies both numerator and denominator. The numerator is the product of the VA non-response adjustment, the under-inclusivity adjustment, and the imputed composite CPHS weight for each decedent, summed over decedents who die in a month. The denominator is the sum of composite CPHS weights in the CPHS sample of living persons that month.

We do not add the numerator (which captures deaths in the population) to this sum because CPHS’s estimate of the population, and thus its inverse probability weights, are based on a projection about India’s population that does not include an adjustment for the pandemic. That is, the weights assume pre-pandemic trends in mortality rates and no additional deaths during the pandemic. This means that our estimate of population-death rates produces an overestimate of death rates: the numerator includes predicted mortality from pre-pandemic trends plus the excess deaths during the pandemic, while the denominator includes living persons and excess deaths during the pandemic but not predicted mortality from pre-pandemic trends. However, adding all pandemic-period deaths to the denominator would lead to an underestimate of the death rate. While it would offset the omission of predicted pre-pandemic mortality in the denominator, it would add pandemic excess deaths a second time, leading to a double-counting of those excess deaths. Although both biases are likely small because death rates are low, we judge that an overestimate is more conservative.

The analogous <u>cause-specific death rates</u> use only VA sample members with the designated cause of death in the numerator.

### Share of deaths attributable to a cause

The <u>share of deaths that are due to a given cause</u> in a month is estimated as the ratio of (a) the cause-specific fully-adjusted death rate in a month and (b) the all-cause fully-adjusted death rate in a month.

### Excess-death rate

The <u>population excess death rate attributable to a cause</u> is defined as the increase in the population death rate from that cause during the COVID pandemic. The numerator of this rate is estimated by subtracting (a) the average fully adjusted death rate from that cause in 2018-2019 from (b) the fully adjusted death rate from that cause during the pandemic. The denominator of that rate is estimated by subtracting (a) the average fully-adjusted all-cause population death rate in 2018-2019 from (b) the fully-adjusted all-cause death rate during the pandemic.

### Heterogeneity in death rates

We estimate variation in SARS-CoV-2 and non-SARS-CoV-2 death rates by attributes such as individual demographics, household geography, and household income via linear regression analysis. We employ a sample of all living CPHS respondents and VA decedents in the period 2020-2021. Data are monthly. A person is recorded as having died in a month by an indicator variable.

We regress an indicator for whether the sample member died in a month from a cause, i.e., was in the VA sample and died in that month due to the cause, against indicators for the decedent’s demographics, locations, and/or household income. Decedent demographics are captured by indicators for 20-year wide age bins and for their sex. Location is captured with an indicator if the household resides in a rural village (using the 2011 Indian Census’ definitions for village).

Household income is measured by the local income quartile to which the decedent’s family belonged in 2017. That indicator is generated using the total household income for each CPHS sample member in all of 2017 to determine quartiles of household income for local areas defined by state and urban status. We include weights that make the regression population-representative. We report regression results that include one set of factors at a time, not all factors at once, so that no category, e.g., females or rural, is omitted for each set. However, we examine a regression with all sets of factors included at once to check if, e.g., the correlation of income on death rates is not driven by age. We multiply coefficients by 12 to ensure that they represent annual rather than monthly death rates.

We determine whether demographic, geographic and income factor have a different correlation with SARS-CoV-2 versus non-SARS-CoV-2 death rates by estimating a regression where the dependent variable is whether the sample member died (regardless of cause) on factors, an indicator for whether the cause of death was SARS-CoV-2, and the interaction of factors with an indicator for whether SARS-CoV-2 was the cause of death. Furthermore, we include weights that make the regression population-representative and multiply coefficients by 12 to ensure that they represent annual rather than monthly death rates.

We show further geographical correlates of SARS-CoV-2 death rates by calculating annual SARS-CoV-2 death rates for each Indian state for the period 2020-2021 and plotting these using a chloropeth map of India.

## III. Results

### Sample size and response rates

#### Living sample

The sample of (living) persons who ever respond to the CPHS contains 892,905 unique persons across 174,003 unique households in the period 2018 to 2021. Because individuals and households rotate into and out of the sample, this amounts to 645,050 unique persons across 131,200 households on average per round in 2018-2021.

Figure 1 plots the number of living individuals (blue line) who respond to a CPHS survey each month from 2018-2021. The household-level response rate to the CPHS survey averaged 85.0% in 2018-2019. That rate fell in March 2020, when India declared a lockdown. It reached a perigee in April 2020 at 30.9% before the peak of India’s first COVID Wave. It substantially recovered between India’s first 2 waves, but briefly dropped again, to a low of 46.1% in May 2021, just after the peak of the second wave. The green line presents the sum of composite CPHS weights, which account for sampling probabilities and CPHS non-response rates, and thus the total population represented by the CPHS sample. This population is linearly increasing and in the range of 1.4 billon persons during the sample period.

**Figure 1.**
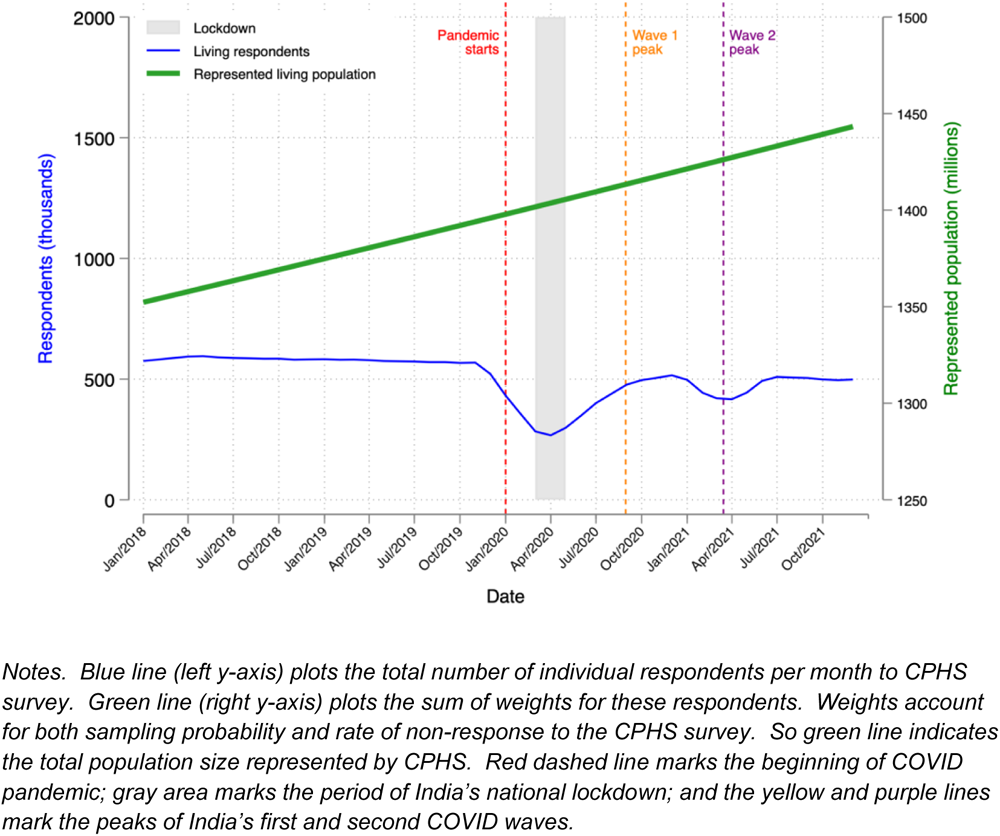
Living CPHS respondents and represented living population by month.

### Deceased sample

The CPHS decedent sample contains 30,447 unique individuals that are reported between 2018-2021 as having died. These individuals appear in the CPHS until the date that their death is reported by living household members in the CPHS survey.

Although VA surveys were attempted for each of these decedents, only 22,178 surveys were completed and included in the actual VA sample, a response rate of 72.8%. Non-response to the VA is not random. Households associated with the CPHS sample of deceased persons that respond to the VA are more likely to, among other things, live in a city (p=<0.001), have fewer household members (p<0.001), have more children (p<0.001), have fewer adult males (p<0.001), not be from a backward caste (p<0.001), self-report poorer health (p<0.001), and not be Hindu (p=0.002) or Muslim (p<0.001) (Supplement, Table S1, column 1). Using propensity score weights (based on the model in column 1, but replacing the rural indicator with strata fixed effect) eliminates these imbalances between VA-responsive and non-responsive households (Supplement, Table S1, column 3).

The CPHS decedent sample is over-inclusive, i.e., includes some deaths that occurred prior to 2018 but were reported after 2018. Among decedents for which a VA was completed, 1,719 (7.8%) had dates of death prior to 2018. Our actual VA sample, which excludes these persons, is 20,453.

The CPHS is also under-inclusive because deaths are reported to the CPHS some months after they occur, a date measured in the VA survey (Supplement, Figure S1). Our adjustment for under-inclusivity accounts for 32-month window of reporting following the date of death. Of the deaths that are reported in this window, 52.7% are reported to CPHS within 4 months of occurring, a lag equal to the period between CPHS surveys of each household; the rate of reporting levels off so that it takes nearly 28 months to reach 98.3% reporting in the 31st month.

Figure 2 plots the number of deaths (orange line) in the actual VA sample that occurred each month. Our use of propensity score weights to correct for VA non-response (green line) increases our estimate of the total number of deaths reported in the CPHS by 39% in the median month. Our adjustment for the delay in reporting deaths to the CPHS survey (i.e., under-inclusivity) increases our estimate of the number of deaths – not just reported deaths – in the CPHS sample by 17% in the median month of the sample period, but by 10.6 times on average in the last 4 months of that period. That spike may be an early indicator of India’s third wave (the Omicron variant), which peaked in January 2022. The purple line reports the sum of weights for each death, thus the estimated total number of deaths in the population (right y-axis) from which CPHS samples, accounting for sampling probabilities and CPHS non-response rates.

**Figure 2.**
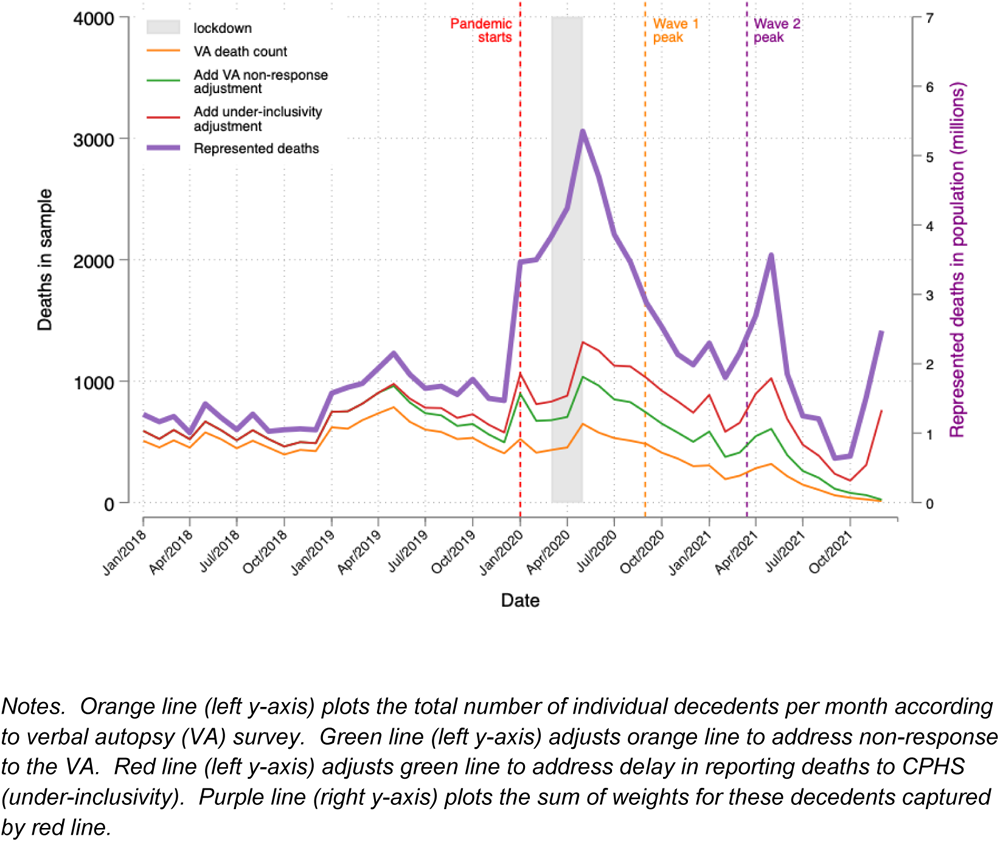
Counts of deaths by month, unadjusted in VA sample, adjusted for VA non-response, adjusted for under-inclusivity of CPHS, and adjusted to be population representative.

### Cause-specific death count

Figure 3A plots our estimate of the number of deaths in the CPHS living sample over time that are attributable to SARS-CoV-2, the 5 leading causes of death prior to the pandemic (cardiovascular diseases, accidents, respiratory disease, diarrheal disease, and malaria), and deaths from other causes. These counts adjust for non-response to the VA survey and delayed or under-inclusive reporting in the CPHS, but not weights that would make them representative of deaths in the population. The count of SARS-CoV-2 deaths is non-zero in 2018-2019. However, we code both U07 and U08 ICD10 codes as SARS-CoV-2-related, even though U07 deaths include vaping-related disorders, which predate the pandemic.

**Figure 3.**
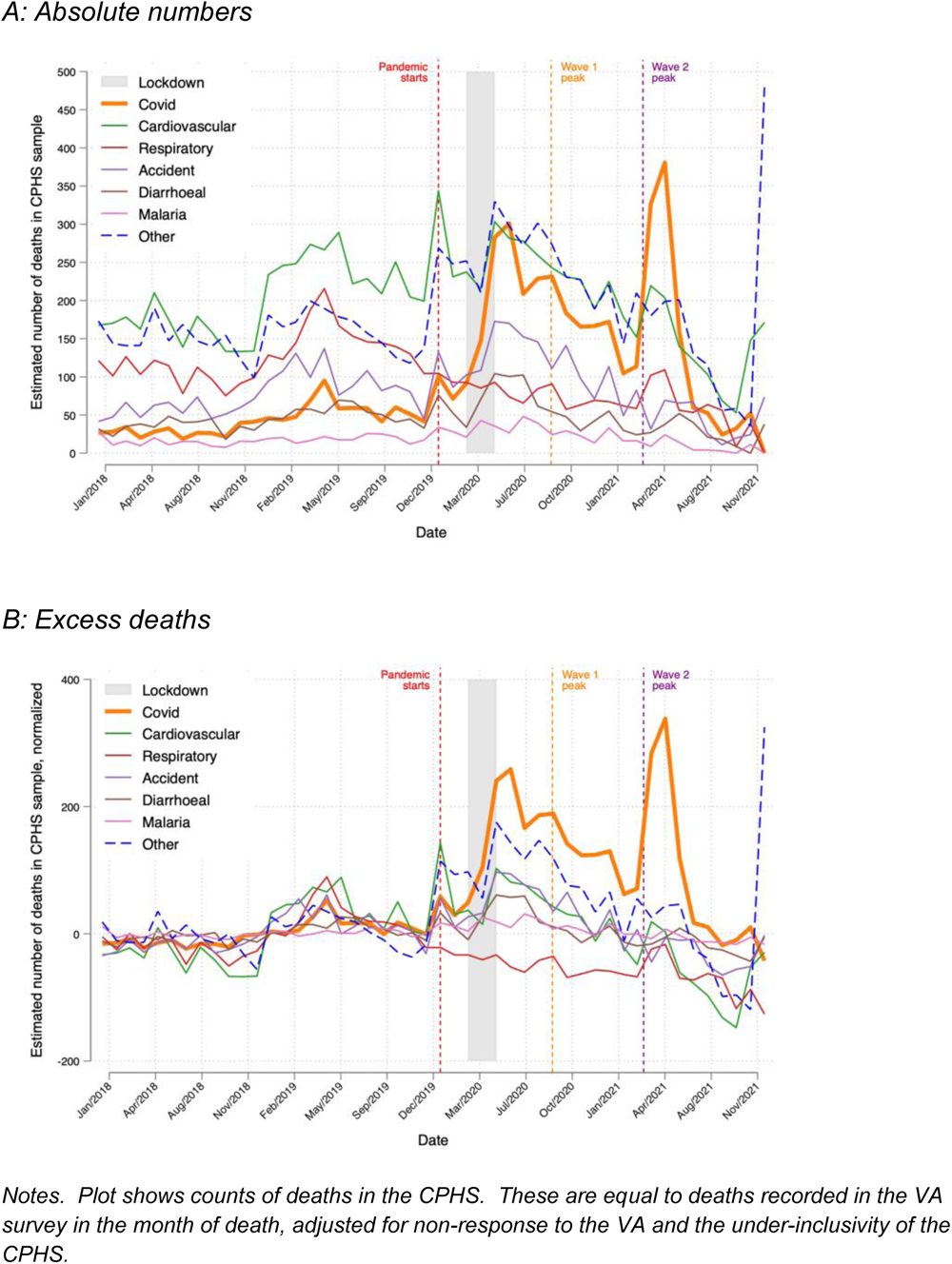
Monthly death counts for SARS-CoV-2, 5 other leading causes of death, and other deaths, 2018-2021.

During the pandemic, deaths coded as U07 or U08 (thus likely SARS-CoV-2) in the CPHS sample spike to (a) to 283 during the lockdown (gray region) and peak just after it, (b) to 231 during India’s first wave (orange vertical line), and (c) to 380 just after India’s second wave (purple vertical line). Cardiovascular and non-specific “other” deaths also spike at the start of 2020 and during the lockdown, around the two COVID waves, and especially at the end of 2021.

Figure 3B reports excess deaths by cause defined as deaths by cause from panel A minus the average number of deaths from that cause in 2018-2019. This highlights the causes of deaths that rise the most during the pandemic. SARS-CoV-2 excess deaths spike more than excess deaths from any other cause during and just after the lockdown (240 deaths), during wave 1 (188 deaths), and just after wave 2 (338 deaths). Respiratory excess deaths fall by 57.4 per month (p<0.001) on average during the pandemic. Due to our under-inclusivity adjustment, “other” cause excess deaths spike at the end of 2021, perhaps related to India’s third COVID wave in early 2020.

### All-cause and cause-specific death rate

Figure 4 plots the monthly all-cause death rate. The orange line gives the raw death rate. The green line adjusts for VA non-response and CPHS under-inclusvity. The red line adds CPHS sampling weights to estimate the death rate in the population. Adjustments have little effect prior to pandemic. The green and red line are similar, suggesting that CPHS sampling probabilities matter less to pandemic death rates than VA non-response rates and CPHS under-inclusivity. Because under-inclusivity weights are large mainly in the last 4 months of 2021, the VA non-response weights have the largest influence on pandemic period death rates. Looking at the red line, the estimated population death rate spikes in January 2020, during the lockdown in May 2020, May 2021 and December 2021 to annualized rates of 3.0, 4.6, 3.0, and 2.1%, respectively.

**Figure 4.**
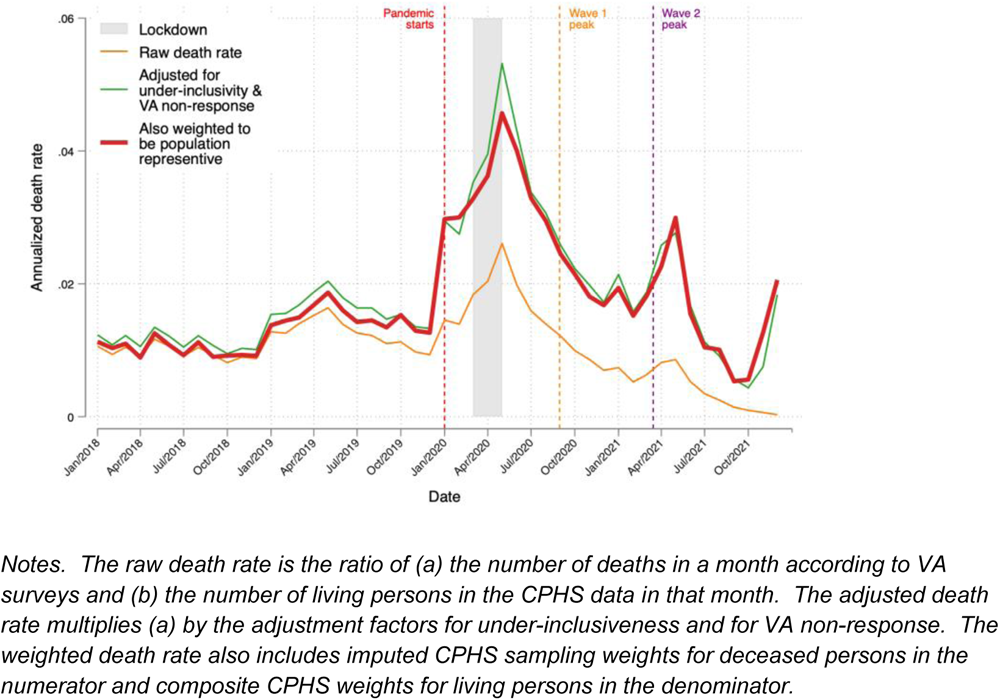
Monthly, all-cause death rate, 2018-2020.

Figure 5A plots cause-specific estimated population death rates for SARS-CoV-2, 5 leading causes of death prior to the pandemic, and other causes of death. During the pandemic, the SARS-CoV-2 death rate peaks at annualized rates of 0.8% in May 2020 during lockdown, 0.9% in June 2020 just before the peak of wave 1, and 1.1% in May 2021 just after the peak of wave 2. These peaks are not proportional to the estimated number of deaths in the CPHS sample because sample members who died at different times may have had different sampling weights. Moreover, the weight of the denominator for the death rate rises as the population of India grows over time. Cardiovascular death rates spike to 1.1% in January 2020, 1.2% in May 2020, and 0.7% in November 2021. Death rates for “other” causes spike to 1.2% in May 2020 and 1.4% in December 2021. Although respiratory deaths decline during the pandemic, respiratory death rates are not significantly lower during the pandemic.

**Figure 5.**
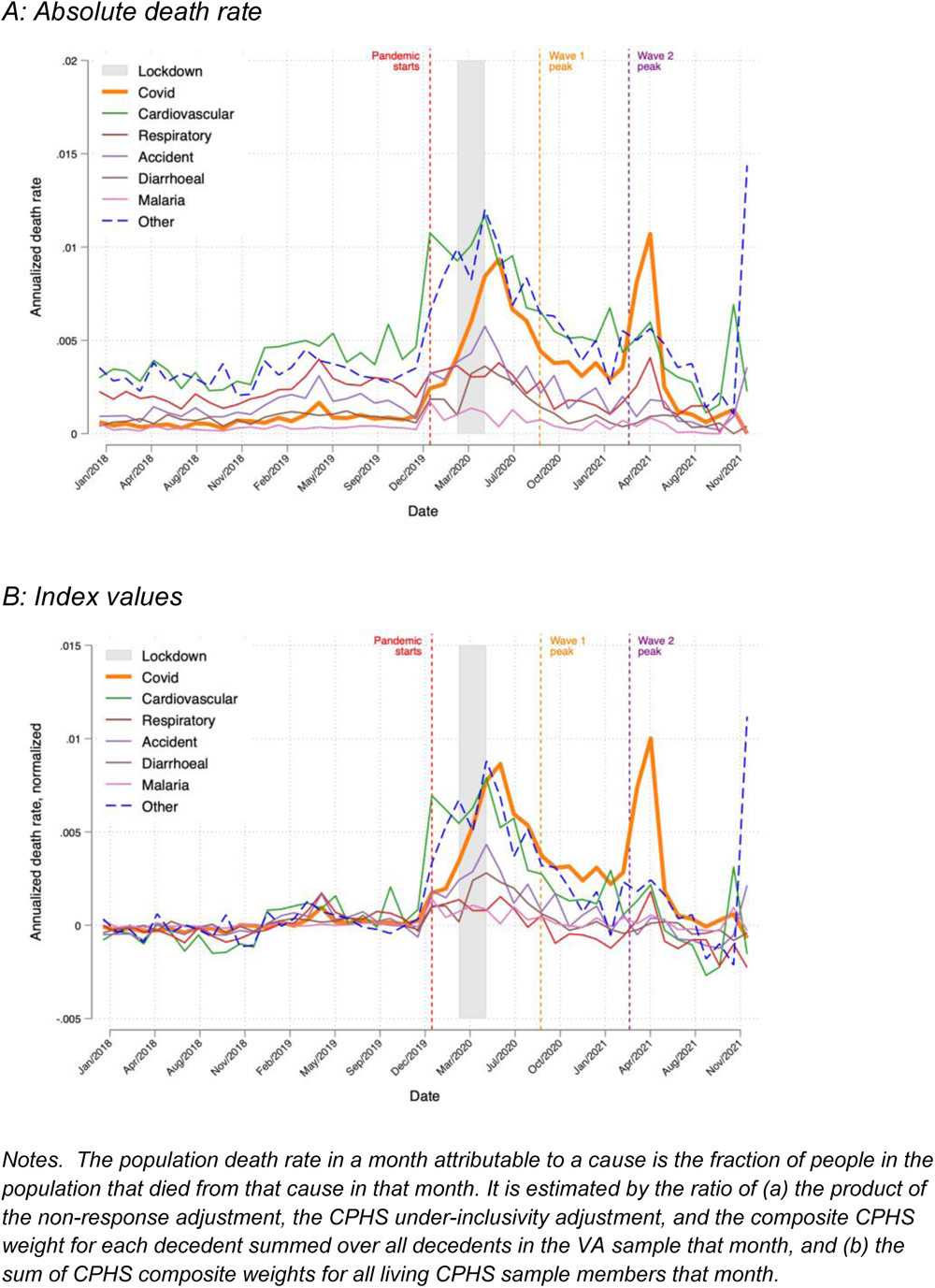
Monthly population death rates by cause of death for SARS-CoV-2, top 5 non-SARS-CoV-2 causes of death, and other deaths, 2018-2021.

Figure 5B plots cause-specific death rates but as an index value, created by dividing cause specific death rates by average death rate for that cause in 2018-2019. One purpose of this index is to net out the effect of vaping-related disorders in the U07 and U08 ICD10 codes so that those codes place greater weight on SARS-CoV-2.

These index values highlight that the spike in death rates was more prominent for SARS-CoV-2 than for cardiovascular or other causes just after the lockdown and especially after wave 2. Moreover, the figure shows that cardiovascular and other cause death rates do not spike after the first few months of the pandemic. After the lockdown they tend to decline. An exception is the last to months of 2021, but the correction for under-inclusivity is driving the spikes. Without that very large adjustment, the spike at the end of 2021 vanishes.

### Share of deaths due to SARS-CoV-2

SARS-CoV-2 becomes a large portion of all deaths starting in 2020 (Figure 6). Its relative importance grows steadily until it spikes in June 2020 and April 2021, when it accounts for over 23.3% and for nearly 35.8% of deaths. After May 2021, the share of deaths attributable to SARS-CoV-2 diminishes.

**Figure 6.**
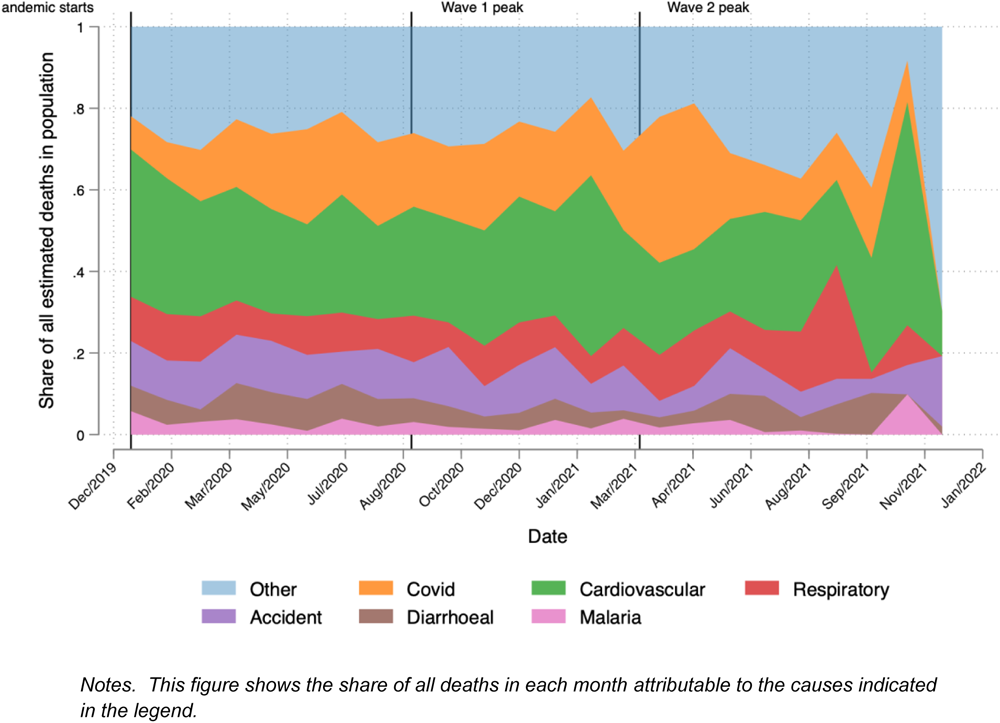
Share of death attributable to SARS-CoV-2 and 5 other top causes of death.

The all-cause annual death rate in 2020-2021 is 81.19% higher than the annual death rate in 2018-2019 (Table 1). SARS-CoV-2 is associated with 17.93% of all deaths during pandemic, and 33.15% of excess deaths during the pandemic. Cardiovascular disease is associated with 27.18% of all deaths and 23.15% of the increase in all-cause death rates during the pandemic. Respiratory disease-related deaths fall by an amount equal to 1.52% of excess deaths during the pandemic. Finally, accidents and tuberculosis are responsible for 8.74 and 5.32% of excess deaths during the pandemic.

**Table 1.** Cause-specific death and excess death rates.

| Cause | Death rate |  | Excess death in 2020-21 |  | Contribution to 2020-21 all cause |  |
| --- | --- | --- | --- | --- | --- | --- |
|  | (1)<br>2018-19 (%) | (2)<br>2020-21 (%) | (3)<br>(pp change) | (4)<br>(% change) | (5)<br>Death rate (%) | (6)<br>Excess death rt (%) |
| All | 1.25 | 2.26 | 1.01 | 81.19 | 100.00 | 100.00 |
| COVID | 0.07 | 0.41 | 0.34 | 484.06 | 17.93 | 33.15 |
| Cardiovascular | 0.38 | 0.61 | 0.23 | 61.75 | 27.18 | 23.15 |
| Respiratory | 0.23 | 0.21 | -0.02 | -6.83 | 9.32 | -1.52 |
| Tuberculosis | 0.04 | 0.09 | 0.05 | 131.17 | 4.13 | 5.23 |
| Neoplasm | 0.06 | 0.07 | 0.01 | 12.03 | 3.20 | 0.77 |
| Ill-defined | 0.04 | 0.05 | 0.02 | 47.87 | 2.36 | 1.70 |
| Digestive | 0.05 | 0.09 | 0.03 | 63.07 | 3.85 | 3.32 |
| Diarrheal | 0.08 | 0.13 | 0.04 | 55.02 | 5.57 | 4.41 |
| Accidents | 0.14 | 0.23 | 0.09 | 62.18 | 10.22 | 8.74 |
| Self-harm | 0.02 | 0.03 | 0.02 | 79.70 | 1.53 | 1.51 |
| Malaria | 0.03 | 0.06 | 0.03 | 104.72 | 2.69 | 3.07 |
| Hepatitis | 0.04 | 0.05 | 0.02 | 41.44 | 2.27 | 1.49 |
| Genitourinary | 0.04 | 0.07 | 0.03 | 62.94 | 2.97 | 2.56 |
| Unknown fever | 0.02 | 0.03 | 0.01 | 67.40 | 1.13 | 1.02 |
| Diabetes/endocrine | 0.02 | 0.04 | 0.02 | 75.07 | 1.68 | 1.61 |
| Epilepsy | 0.01 | 0.02 | 0.01 | 104.83 | 0.76 | 0.87 |
| Sexual diseases | 0.00 | 0.00 | 0.00 | -84.39 | 0.02 | -0.18 |
| Preventable by vaccine | 0.00 | 0.00 | 0.00 | 138.57 | 0.11 | 0.15 |
| Meningitis/encephalitis | 0.01 | 0.01 | 0.00 | 28.66 | 0.33 | 0.17 |
| Maternal | 0.00 | 0.01 | 0.00 | 246.62 | 0.26 | 0.41 |
| Other infectious diseases | 0.03 | 0.04 | 0.01 | 31.71 | 1.94 | 1.04 |

Note. Columns 1 and 2 provide the death rate attributable to various causes and all causes in 2018-2019 (pre-pandemic) and 2020-2021. Columns 3 and 4 provide the excess cause-specific death rate in percentage points (pp) and percent of 2018-2019 same-cause death rate. Columns 5 and 6 provide the contribution each cause made to the 2020-21 all-cause death rate or excess death rate.

### SARS-CoV-2 death rate by demographic, geographic and economic group

The death rate from SARS-CoV-2 is significantly higher in higher age groups. It is highest among individuals who were above the age of 80 when they died, followed by those between ages 61-80 (Figure 7a, blue dots). However, because so few people are over 80, the vast bulk of SARS-CoV-2 deaths were among those between ages 61-80 (blue bars). Death rates from non-COVID causes are also significantly higher in higher age groups (Figure 7b, blue dots). Indeed, the age gradient on non-COVID death rates is significantly greater than the age gradient on SARS-CoV-2 death rates (p < 0.001).

**Figure 7.**
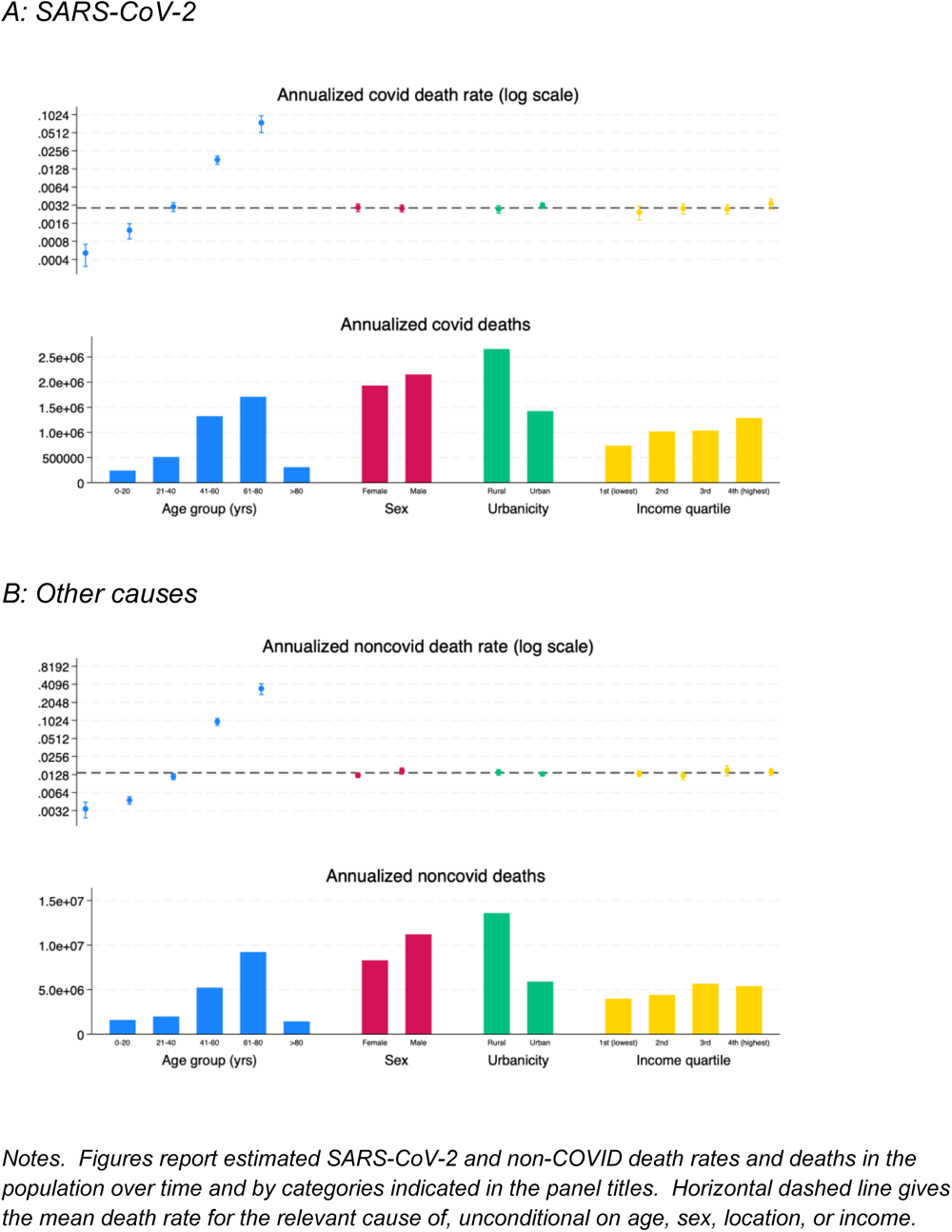
Death rates and deaths from SARS-CoV-2 (panel A) and other causes during the pandemic, by age group and by rural or urban residence (panel B).

The SARS-CoV-2 death rate is higher among females, among urban residents, and in the highest income quartile, but not significantly so (Figure 7a). While urban residents have higher death rates, total deaths are greater in rural areas because there are more residents in rural areas. Non-COVID death rates follow different patterns than SARS-CoV-2 (Figure 7b). They are higher among males and rural residents, though not significantly so. The differences between non-COVID and SARS-CoV-2 death rates are significantly different for rural residents, but not for males or different income quartiles.

Figure 8 presents the annual SARS-CoV-2 excess death rates for different states during the pandemic. Death rates tend to be lower in the Gangetic plain (e.g., Uttar Pradesh 0.13%; Bihar 0.07%; Jharkand, 0.07%) and highest in Goa (2.13%), Meghalaya (1.8%), and Tripura (1.42%).

**Figure 8.**
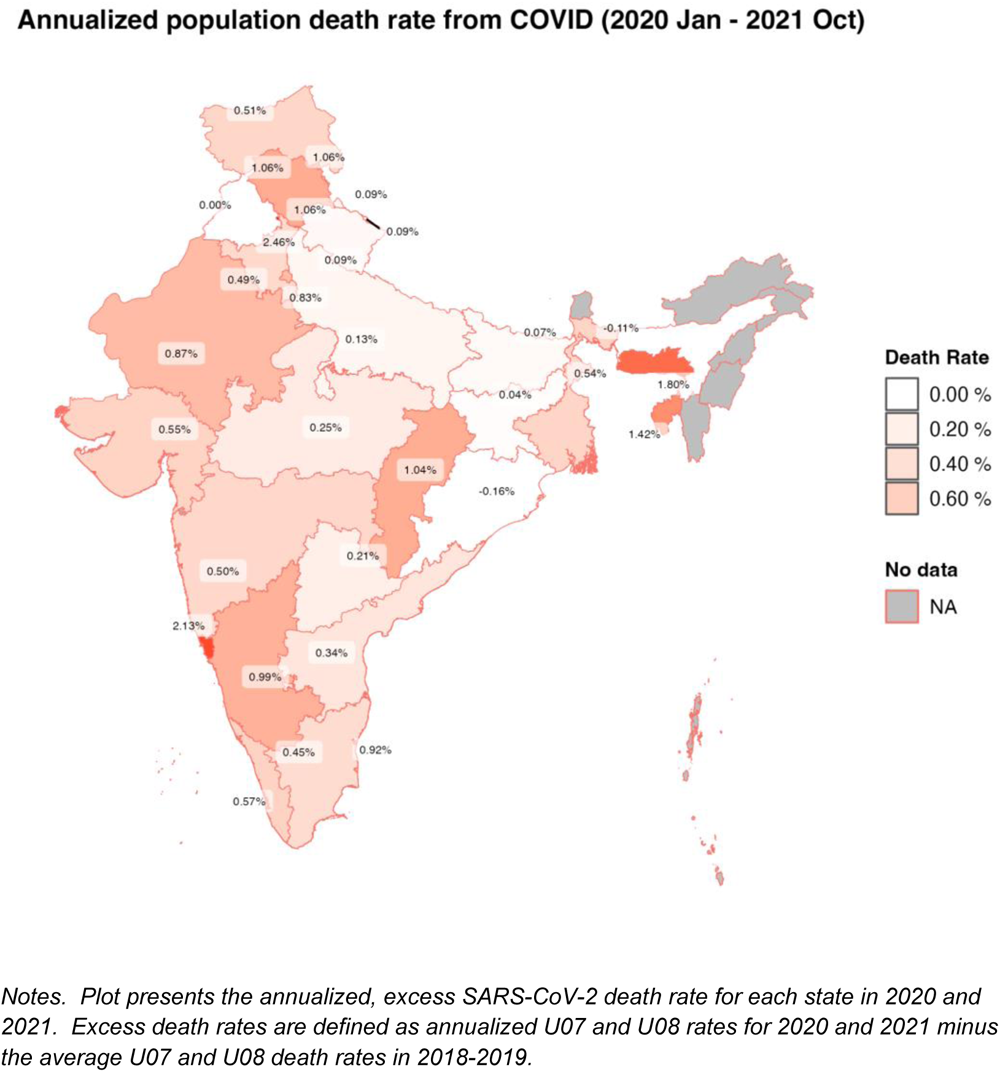
Excess SARS-CoV-2 death rates by state during the pandemic.

## Discussion

We completed verbal autopsy (VA) surveys on a sample of 20,453 persons who are reported in 2018-2020 as having died to determine the cause of death. VA surveys were attempted on a representative sample of 30,447 deaths in the CPHS sample during this period. The final VA sample reflects a 72.8% response rate on attempted surveys, plus the elimination of 7.8% of completed surveys because the deaths actually occurred prior to 2018.

The timing of spikes in COVID waves differs somewhat from official reports. Official reports of SARS-CoV-2-attributed deaths spike during India’s first (September 2020) and second (April 2021) waves. VA surveys confirm the spike during wave 2. However, the 2020 spike occurs at the tail end of the lockdown (May-June 2020) rather than during wave 1. Our findings are consistent with the finding that SARS-CoV-2 rates spiked during the lockdown, especially among the urban poor ^18^.

Death rates from SARS-CoV-2 disproportionately spike only during wave 2. SARS-CoV-2 death rates spike at the end of the lockdown and during wave 2. However, death rates from other causes – notably cardiovascular disease and all “other” causes also spike during the lockdown. This gap between SARS-CoV-2 and non-COVID death rates may explain why wave 2 seemed especially stark in India.

Death rates do not translate directly into deaths. SARS-CoV-2 death rates increase significantly with age, with populations above age 80 having the highest death rates. However, there were fewer deaths of those above 80 because there are far few people above 80 than between 60 and 80 in the population. Likewise, while SARS-CoV-2 death rates were greater in urban areas than in non-urban areas, there were more SARS-CoV-2 deaths in rural areas because the rural population is larger than the urban population.

SARS-CoV-2 deaths are surprisingly less correlated with age than non-COVID deaths. SARS-CoV-2 death rates increase significantly with age, with populations above age 80 having the highest death rates. However, non-COVID death rates also increase with age. Indeed, the difference between the over-80 death rate and the under 20 death rate is significantly greater for non-COVID causes of death than for SARS-CoV-2 (p<0.0001).

Income does not have a clear an impact on SARS-CoV-2 deaths. While ^6^ finds that all-cause excess death rates are significantly, positively correlated with income during the pandemic, we find that the correlation between the SARS-CoV-2 death rate and income is not significant.

SARS-CoV-2 explains 17.93% of deaths and 33.15% of excess deaths during the pandemic. Cardiovascular deaths are responsible for 23.15% of excess deaths during the pandemic. However, some of these individuals may have died in part due to SARS-CoV-2. Individuals with cardiovascular disease are at greater risk of mortality conditional on SARS-CoV-2 infection ^19^. Moreover, SARS-CoV-2 may cause cardiac damage ^20,21^. So, it is possible SARS-CoV-2 deaths are underestimates by the VA survey.

Respiratory deaths fell during the pandemic, by an amount equal to 1.52% of excess deaths. It is possible that some of the increase in SARS-CoV-2 deaths are actually mis-coded respiratory deaths. That suggests that we may overestimate SARS-CoV-2 excess deaths, though the magnitude of the overestimate is small.

This paper has several limitations. The VA survey suggests there were death attributable to ICD10 categories U07 and U08 in 2018-2019. While we call U07 and U08 deaths as SARS-CoV-2 deaths, U07 includes deaths associated with vaping related disorders. This may be why Figure 3A reports a positive number of SARS-CoV-2-labeled deaths in 2018-2019. Vaping was a public health crisis prior to the pandemic ^22^, including in India ^23^. We attempt to correct for this by converting cause-specific death rates into index values in Figure 5B and by calculating excess death rates in Table 1: both these steps net out average death rates in 2018-2019. However, vaping may remain partly responsible for the rise in excess SARS-CoV-2-related deaths in 2020-2021 because vaping may compound the risk from SARS-CoV-2 infection ^22^. In general, one must be cautious about relying on mono-causal explanations of death.

Second, the completed VA sample requires statistical corrections to be considered representative, but those corrections may be imperfect. The representative sample of CPHS deaths that is used as the sample frame for the VA is missing deaths which occur during between 2018-2021 but are not yet reported to CPHS. We correct that with a factor that accounts for the statistical delay in reporting deaths. But that correction can only correct for deaths that are ultimately reported in 48 months; we have no information and cannot correct for deaths that are reported more than 4 years after occurring. Moreover, our correction lead to surprisingly sharp increases in death rates from “other” causes, which are hard to understand.

In addition, the households that respond to the VA survey are not a random sample of all CPHS households that reported a death in 2018-2021. We correct for this with a propensity score weight, which does a reasonable job eliminating imbalance in attributed between households that do and do not respond to VA surveys. However, our adjustment cannot correct for imbalance on variables that are not used in our propensity score estimation model.

Third, the verbal autopsy survey we employed was well-validated and used prior to the pandemic ^12,13^. However, the procedure for mapping answers to diagnostic codes had to be modified for a new diagnosis, SARS-CoV-2. The specific modification we employed has not yet been validated – although other modifications to the WHO VA instrument to address COVID have been ^24,25^. Nevertheless, assessments of VA modifications to capture SARS-CoV-2 show there is error and diagnosis should be complemented with minimally invasive techniques such as ultrasound when the decedent’s cadaver is available ^26^.

## Data Availability

All data in the present study will be made available to the public via the open data website openmortality.org.

https://consumerpyramidsdx.cmie.com/

## Acknowledgements

We acknowledge the critical support of Prabhat Jha and Leslie Newcombe from the Centre for Global Health Research and Mahesh Vyas at the Centre for Monitoring Indian Economy (CMIE). CMIE conducted the verbal autopsy surveys. Jha and Newcombe, along with their team at CGHR, mapped answers to the survey onto ICD10 codes. Jha and Newcombe also helped provide us cleaned data on date of death and cause of death. We also thank Emergent Ventures for funding the creation of the CMIE Verbal Autopsy database.

## Supplement

### I. Methods

#### CPHS

##### Strata

CPHS samples randomly within strata. Strata are defined in two dimensions. One dimension is a cluster of similar districts within a state which CPHS calls a homogenous region. The second dimension is 5 categories of human settlement type and size (very large towns with over 200,000 households, large towns with 60,000 - 2000,000 households, medium towns with 20,000-60,000 households, small towns with less than 20,000 households, and rural villages that the Indian Census distinguishes from towns). The last of these categories is called a rural strata; the remaining are called urban strata. In 2018-2021, there were 102 unique homogenous regions and 382 unique homogenous region x town size strata.

##### Sampling in strata

CPHS samples homes in differently in rural and urban strata ^27^. In each rural strata, CPHS randomly selects roughly 30 villages in that strata. The number of villages selected per strata grows over time, but weights are adjusted to account for changes in this count and ensure that estimates from the sample can be made population-representative.

Within each chosen village, CPHS chooses a random starting point on the main street of the village and then engages in systematic sampling, attempting to survey every n-th house on that street until it is able to sample 16 houses in the village. The value of n ranges from 5 to 15 and is at picked at random.

In each urban strata that exists, CPHS randomly picks at least one town in the strata. (Some larger town sizes are not present in some homogenous regions.) Again, the number of urban strata and towns selected grows over time, but weights are adjusted to ensure estimates from the sample can be made population representative. In each selected town CPHS randomly selects 21 Census Enumeration Blocks, each a cluster of 100-125 households defined by the Indian government for the purpose of its decennial census. Finally, within each census block CPHS, CPHS uses systematic sampling to select 16 households.

##### CPHS survey

The CPHS survey creates a roster of each current member of a household. It also records household member-level changes in that roster and the reasons for the change, including birth, death, immigration away from household, and immigration into the household. The sample of deceased individuals in the CPHS data are obtained from this roster.

For each member of the household or the household as a whole, as appropriate, the CPHS survey captures demographics, income, consumption and financial status including assets and debt. Demographics include age and sex, as well as religion and caste. Income data permit calculation of quartile of income distribution.

### Weights for living CPHS sample members

Weights for the living sample in the CPHS are premised on the survey having employed a stratified design with random sampling within strata. A living respondent’s “sampling weight” is N(j,t)/n(j,t), where N(j,t) is the total population in strata j in month t, and n(j,t) is the size of the CPHS sample in strata j at time t. Strata are defined by location, urban status, and town size if location is urban (as explained in the Supplement).

One problem with these sampling weights is that not all CPHS sample households respond to each CPHS survey wave. To address non-response, CPHS provides “non-response weights” for each responding living sample member to account for non-responding living sample members. These non-response weights assume non-response is random within strata. The weight for a responding sample member in strata j at time t is M(j,t)/m(j,t), where M(j,t) is the total CPHS sample intended to be surveyed in strata j in month t and m(j,t) is the number of CPHS respondents in strata j in month t. We multiply the sampling weight by the CPHS non-response weight to obtain the “CPHS weight” of each living respondent in the CPHS in each month.

### Weights for deceased CPHS sample members

#### Adjustment for under-inclusivity of deceased sample

The VA sample only includes deaths reported in CPHS during 2018-2021. Because deaths are reported with a delay, the CPHS sample of deaths is under-inclusive. To address this, we estimate an adjustment factor that we use to scale population all-cause and cause-specific death rates each month from 2018-2020 to capture all deaths that would be reported in 32 months, if 32 months of reports were available. (We chose 32 months because, in our secondary measure of date of death, i.e., midpoint between surveys, no death is reported later than 32 months after it occurs. Moreover, in our primary measure of the date of death, the VA survey, over 98% of deaths reported within 48 months are reported within 32 months.) This adjustment factor for death rates in a given month is the ratio of (a) the number of deaths that would be reported after 32 months of reporting and (b) the number of deaths that would be reported in t months, where t is the number of months between the given month and December 2021, the last date of reporting for this study.

This adjustment factor estimated in three steps. First, we isolate decedents whose date of death is in the first 16 months of the VA sample, i.e., Jan. 2018-April 2019, inclusive. Second, for each month we calculate the ratio of (a) the number of deaths that month that were reported within 32 months of occurrence and (b) the number of deaths that month that were reported within t months, where t ranges from 1 to 32. This results in 32 ratios per month of death, for 16 months. Third, we take averages across the first 16 months of death for each t, result in 32 averages of (a)/(b). These 32 averages are our adjustment factors.

Note that the reciprocal of these adjustment factors give the fraction of deaths that are reported in 32 months that are reported within t months, where t ranges from 1 to 32. We plot these factors, which sketch an empirical cumulative distribution function for time until a death is reported, to illustrate the degree of adjustment required for each t in Figure S1.

### VA non-response weights

We create weights to account for the fact that not all households with a decedent reported to CPHS between 2018-2021 respond to the VA survey. These weights are constructed and evaluated in three steps. First, using the sample of all persons reported to CPHS as deceased from 2018-2021, we estimate a probit regression wherein the dependent variable is an indicator for whether the decedent’s household responded to the VA and the independent variables are a range of household-level variables (number of members, number of children, and average adult age; religion and caste; educational status, literacy, and employment status of head of household; possession of mobile phone; CPHS weights) and strata x rural status fixed effects and report the results as an unadjusted test of non-random VA response. These data are measured as of CPHS round 1 in 2022, when we executed the VA survey, or the first prior month in which we have such data from a household. If a household has multiple deceased members, the household is included multiple times in this regression. We present the results of this regression in column 2 of Table S1 below. (Column 1 replaces the strata x rural fixed effects with a rural dummy to illustrate that rural status correlates with VA non-response.) We report p-values for variables that predict the largest differences in propensity to respond to the VA and the extent of variation in response explained by strata fixed effects.

Second, we estimate predicted probabilities p(i) of VA response for each deceased person i’s household. We calculate propensity-score adjusted weights for each deceased person; these are 1/p(i) for a person whose household responded to the VA and 1/(1-p(i)) for a person whose household did not. We re-run the regression from step one with these weights and report the results as an adjusted test of non-random VA response. To validate our weights, we re-estimate the probit regression in step one, but employing the propensity score-adjusted weight for deceased persons from responding and non-responding households. We present the results of this regression in the third column of Table S1.

Third, we use the propensity score-adjusted weight for deceased persons whose households respond to the VA to adjust for non-response to the VA when calculating all-cause or cause-specific death counts and rates. We do not use the propensity score adjusted weights for deceased persons whose households did not respond to the CPHS because we do not have cause of death for those individuals.

### Alternative dates of death

In the main text, we report results where the date of death is obtained from VA interviews.

When those interviews do not produce a “viable” date of death that is before the date on which a death is reported to CPHS, we use an alternative measure that is the midpoint between when a death is reported to CPHS and the last date that the decedent’s household responded to a CPHS survey. Here we expound on the alternative measure and provide results if the alternative measure were used even when the VA survey provides a viable date of death.

The alternative date of death is premised on three facts: the CPHS survey is attempted on each sample household every four months, a decedent’s family may not be interviewed in the month in which the decedent dies, and the CPHS does not capture the exact date of death. These facts imply that the date on which a decedent is reported as passed to the CPHS, which we call the date of report (DOR), is later than the actual date of death. They also imply that the date of death is after the last time that a household is successfully surveyed by the CPHS, which we call the date of last survey (DOLS).

Because the date of CPHS surveys of a given household are independent of the date of the decedent’s death and CPHS surveys roughly the same number of households per month, our alternative measure of the date of death is the midpoint between DOLS and DOR. If the decedent died between those two surveys, the midpoint is an unbiased estimate of the date of death.

How this is implemented is that if a household responded to two consecutive surveys, then we assign the date of death to be DOR minus 2, because surveys are 4 months apart. If the household skipped a survey, then the midpoint is further back from the DOR. For example, if 1 survey is skipped, then the gap is 8 months, so the midpoint is DOR - 4. In general, the alternative measure is DOR - ((s+1) x 2), where s is the number of surveys the household skipped. The maximum gap between any two surveys in our sample is 64 months, and so all deaths are within 32 months of the DOR.

Figure S2 reports the fraction of deaths that are reported to CPHS within t months (i.e., t months or less) following a death under the alternative dating of deaths. The x-axis is the number of months t since a death occurred. The y-axis is the fraction of deaths captured in the VA survey.

Figure S3A and B reports cause-specific deaths and death rates, respectively, using the alternative coding of the date of death. The new coding of death shows that SARS-CoV-2 is not as large a relative cause of death in 2020-2021. The reason is not that there are fewer SARS-CoV-2 deaths with the new date. Rather, the explanation is that cardiovascular and other deaths that are attributed to 2018-2019 in the dating method used in the main text are pushed “forward” into 2020-2021 in the alternative dating in this Supplement. This can be seen by comparing the average level or rate of cardiovascular and “other” deaths in 2020-2021 compared to 2018-2019. This implies the mid-point between surveys tends to be later than the date of death reported by the decedent’s family.

## II. Results

**Table S1.**
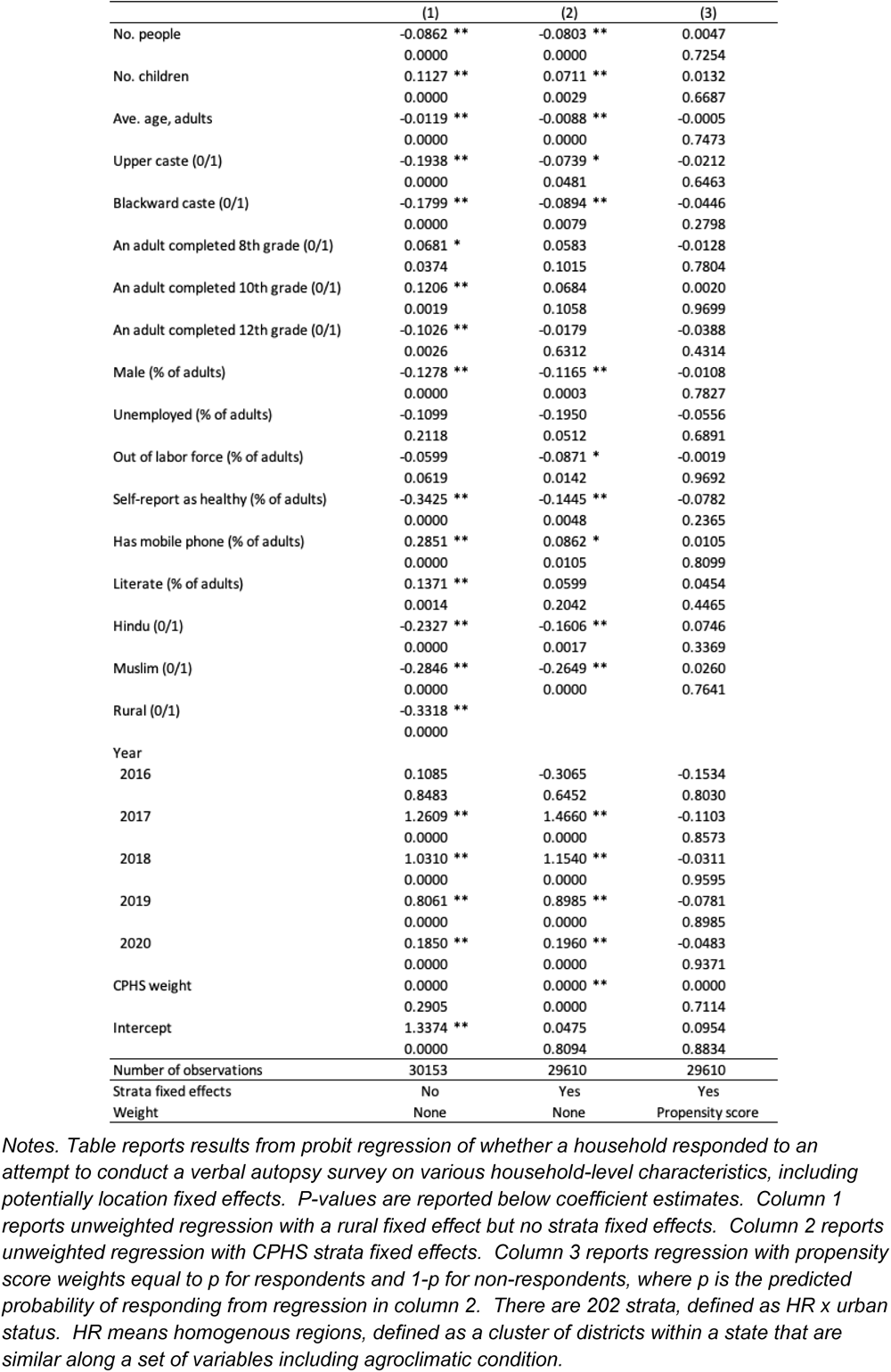
Balancing test and propensity score weight validation.

**Figure S1.**
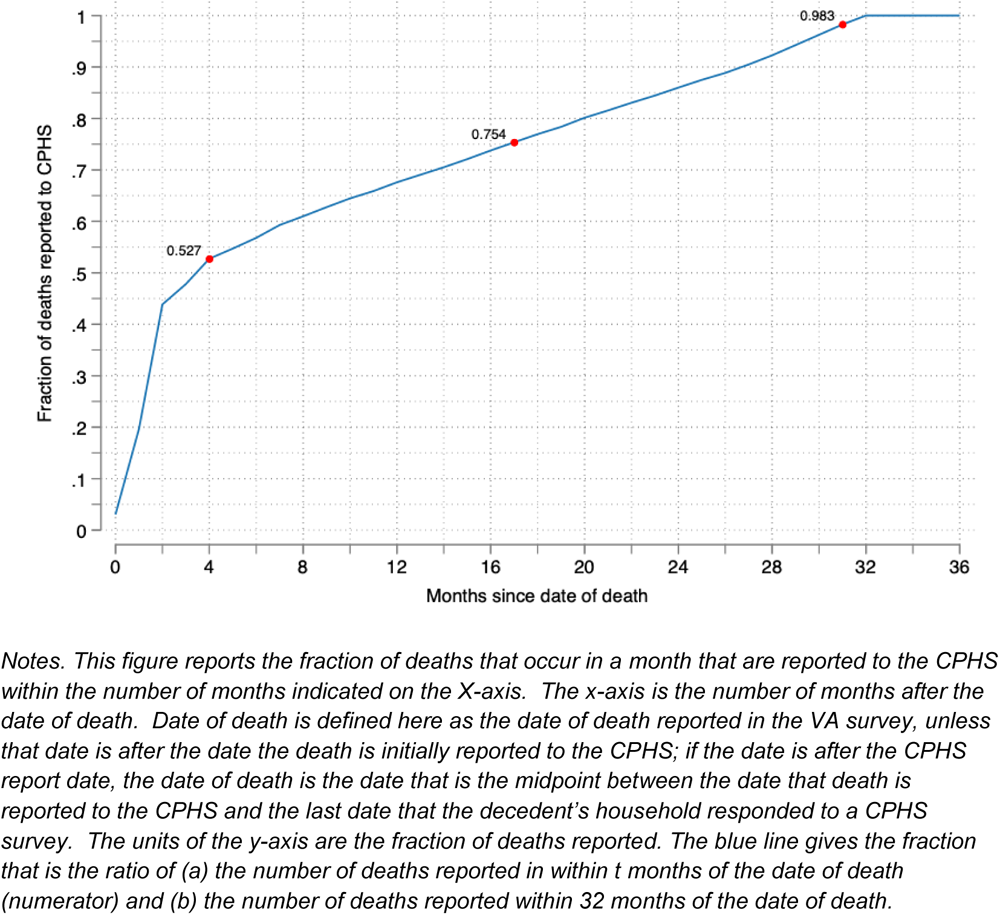
Delay in reporting deaths to the CPHS using date measurement in main text.

**Figure S2.**
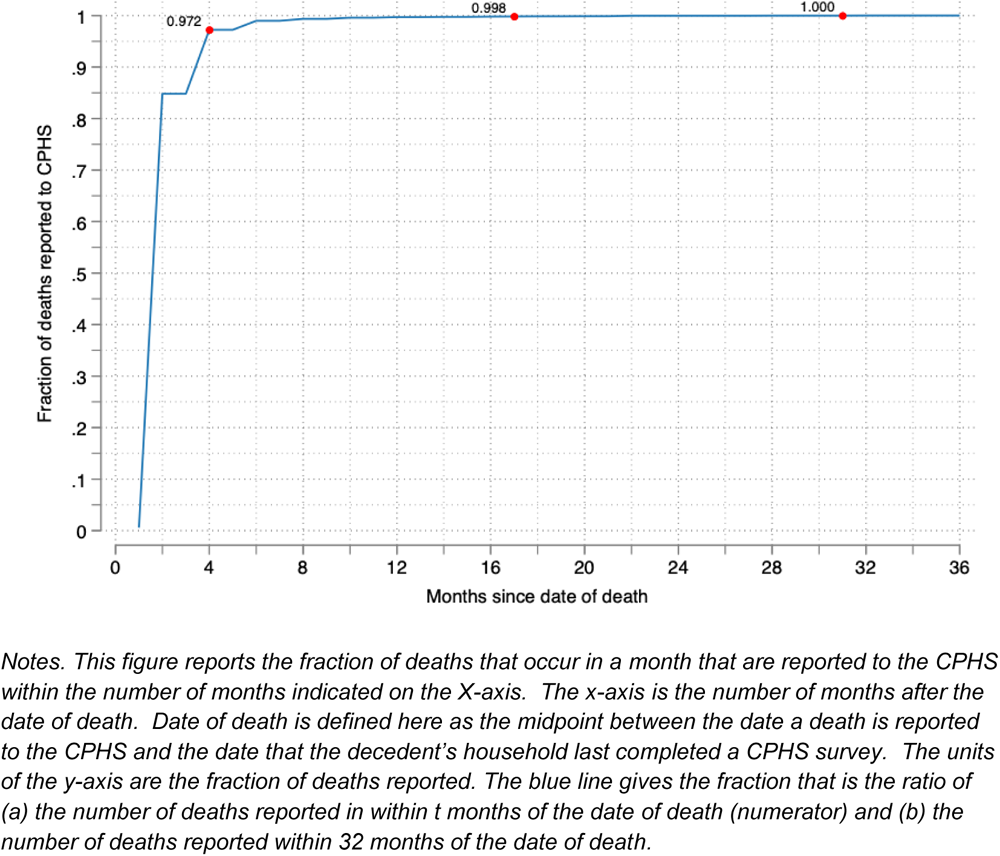
Delay in reporting deaths to the CPHS using alternative date measurement (mid-point between date of report and last completed survey).

**Figure S3.**
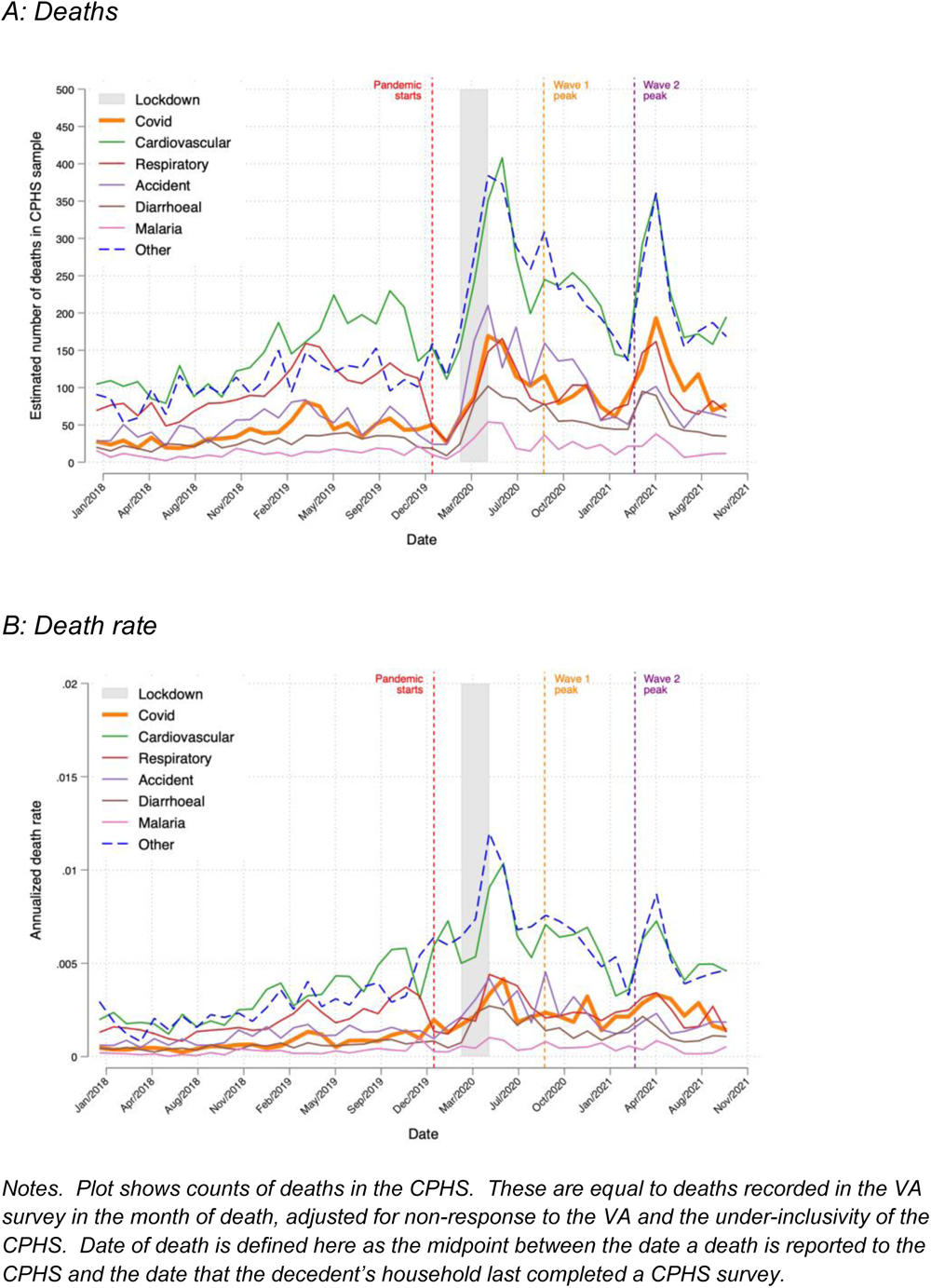
Monthly death counts for SARS-CoV-2, 5 other leading causes of death, and other deaths, 2018-2021, using the alternative date of death.

